# Impaired antigen-specific memory B cell and plasma cell responses including lack of specific IgG upon SARS-CoV-2 BNT162b2 vaccination among Kidney Transplant and Dialysis patients

**DOI:** 10.1101/2021.04.15.21255550

**Authors:** Hector Rincon-Arevalo, Mira Choi, Ana-Luisa Stefanski, Fabian Halleck, Ulrike Weber, Franziska Szelinski, Bernd Jahrsdörfer, Hubert Schrezenmeier, Carolin Ludwig, Arne Sattler, Katja Kotsch, Alexander Potekhin, Yidan Chen, Gerd R. Burmester, Kai-Uwe Eckardt, Gabriela Maria Guerra, Pawel Durek, Frederik Heinrich, Marta Ferreira-Gomes, Andreas Radbruch, Klemens Budde, Andreia C. Lino, Mir-Farzin Mashreghi, Eva Schrezenmeier, Thomas Dörner

## Abstract

Patients with kidney failure are at increased risk during the COVID-19 pandemic and effective vaccinations are needed. It is not known how efficient mRNA vaccines mount B and plasma cell responses in dialysis patients (DP) or kidney transplant recipients (KTR) compared to healthy controls (HC). We studied humoral and B cell responses of 25 HC, 44 DP and 40 KTR. Markedly impaired anti-BNT162b2 responses were identified among KTR and DP compared to 100% seroconversion in HC. In DP, the response was delayed (3-4 weeks after boost) and reduced with anti-S1 IgG positivity in 31 (70.5%) and anti-S1 IgA in 30 (68.2%) of 44, respectively. In contrast, KTR did not develop IgG response except one patient who had prior unrecognized infection and developed anti-S1 IgG. The majority of antigen-specific B cells (RBD+) were identified in the plasmablast or post-switch memory B cell compartments in HC, whereas these RBD+ B cells were enriched among pre-switch and naïve B cells from DP and KTR. Single cell transcriptome and CITE-seq analyses found reduced frequencies of plasmablasts, *TCF7*+*CD27*+*GZMK*+ T cells and proliferating *MKI67*-expressing lymphocytes among KTR non-responders. Importantly, the frequency and absolute number of antigen-specific circulating plasmablasts in the whole cohort correlated with the Ig response, a characteristic not reported for other vaccinations. In conclusion, this data indicate that lack of T cell help related to immunosuppression results in impaired germinal center differentiation of B and plasma cell memory. There is an urgent need to improve vaccination protocols in patients after kidney transplantation or on chronic dialysis.

**One Sentence Summary:** Kidney transplant recipients and dialysis patients show a markedly diminished humoral response and impaired molecular B cell memory formation upon vaccination with BNT162b2.

## INTRODUCTION

The current coronavirus disease 2019 (COVID-19) pandemic leads to a high morbidity and mortality especially among patients with kidney failure (*1*). Both, patients on maintenance dialysis patients (DP) and kidney transplant recipients (KTR) are at increased risk of developing COVID-19 and experiencing a severe course, due to a number of factors, including inevitable exposure risk in the health care system, their co-morbidities and impaired immune function, either as a consequence of kidney failure or immunosuppressive medication after kidney transplantation. For this vulnerable and highly exposed population, vaccination is of utmost importance. The development of effective vaccinations progressed rapidly and different vaccines have been approved.

One of these vaccines is the mRNA SARS-CoV-2 vaccine BNT162b2 (BioNTech/Pfizer) which has demonstrated efficacy in healthy individuals in a clinical study (*2*) and under real-world conditions (*3*). Recent data have described a lower serological response to an mRNA vaccine in dialysis patients (*4*) and kidney transplant recipients (*5*), suggesting an overall diminished vaccine response. Whereas numerous studies have addressed the consequences of conventional vaccines on B and plasma cells (*6-8*) and corresponding Ig levels, nothing is known yet about the B lineage consequences in response to an mRNA vaccine among healthy controls and immunocompromised patients. Previous studies in kidney failure patients reported markedly diminished response to vaccinations. This has led to an adaption of vaccination protocols with either higher initial vaccine doses or more frequent booster doses (*9, 10*). If such adaptations are required for mRNA vaccines is not yet known.

Protection through immunization is achieved by an orchestrated immune response between different cellular subsets of innate (antigen presenting cells (APCs)) and the adaptive immunity, B and T cells. The humoral immune response leads to the production of antibodies by antibody secreting cells (ASCs), which can provide rapid protective immunity. Moreover, the generation of B cell memory, which in case of infection leads to a rapid expansion and production of high affinity antibodies, is a central component of a robust secondary immune response. Both, memory B cells and ASCs have a long lifespan, even without re-exposure to the antigen (*11*). The induction of B cell memory by mRNA vaccines and the relation to humoral immune response is largely under-investigated, including studies of immunocompromised cohorts. Upon natural acute SARS-CoV-2 infection, immunological memory (antibodies and memory B cells) has been shown to last for at least 8 months (*12*). In patients with chronic kidney disease, these data are largely lacking, although prolonged time of viral shedding with impaired virus clearance have been reported likely related to impaired cell-mediated immunity (*13*).

In this study, we compared the characteristics of the humoral and antigen-specific B cell immune response against the mRNA vaccine BNT162b2 between healthy controls and patients with kidney failure treated by maintenance hemodialysis or kidney transplantation. We found a diminished humoral response to BNT162b2, and a lack of proper B lineage memory formation including RBD-specific plasmablasts and of post-switch memory B cells.

## RESULTS

### Cohorts and patient characteristics

For this study, we recruited 25 healthy controls (HC), 40 patients on maintenance hemodialysis, 4 peritoneal dialysis patients and 40 KTR. Hemodialysis and peritoneal dialysis (PD) patients did not significantly differ in age and vaccine response and were therefore grouped together. After written informed consent, serum and peripheral blood mononuclear cells (PBMCs) were collected before vaccination (baseline) and 7±2 days after boost vaccination (second dose), respectively. Serological follow-up was available in DP and KTR patients 3-4 weeks after boost. Due to local vaccination guidelines, HC who were mainly health care workers were significantly younger than DP and KTR (p< 0.01) while DP and KTR had a similar age. Among HC the age group between 60 and 69 is missing due to the current German vaccination guidelines. As known for patients with kidney failure (*14*), a majority of DP and KTR were male. The median time on dialysis was 5.5 years (IQR, 2.0, 9.0). Among KTR only one patient was transplanted less than one year ago and median time after transplantation was 5.0 years (IQR. 2.0, 10.0). KTR were on a uniform immunosuppressive regimen with mycophenolate mofetil (MMF) in 39/40, steroid in 37/40 and calcineurin inhibitor (CNI) in 37/40 patients. Demographics are summarized in Table 1. To identify previously SARS-CoV-2 infected individuals we measured anti-nucleocapsid protein (NCP) antibodies 7±2 days after boost, which is not a component of BNT162b2. Therefore, positivity of NCP originates from natural infection. One HC, one DP and one KTR was identified with positive NCP (supplementary Figure 1).

**Table 1:**
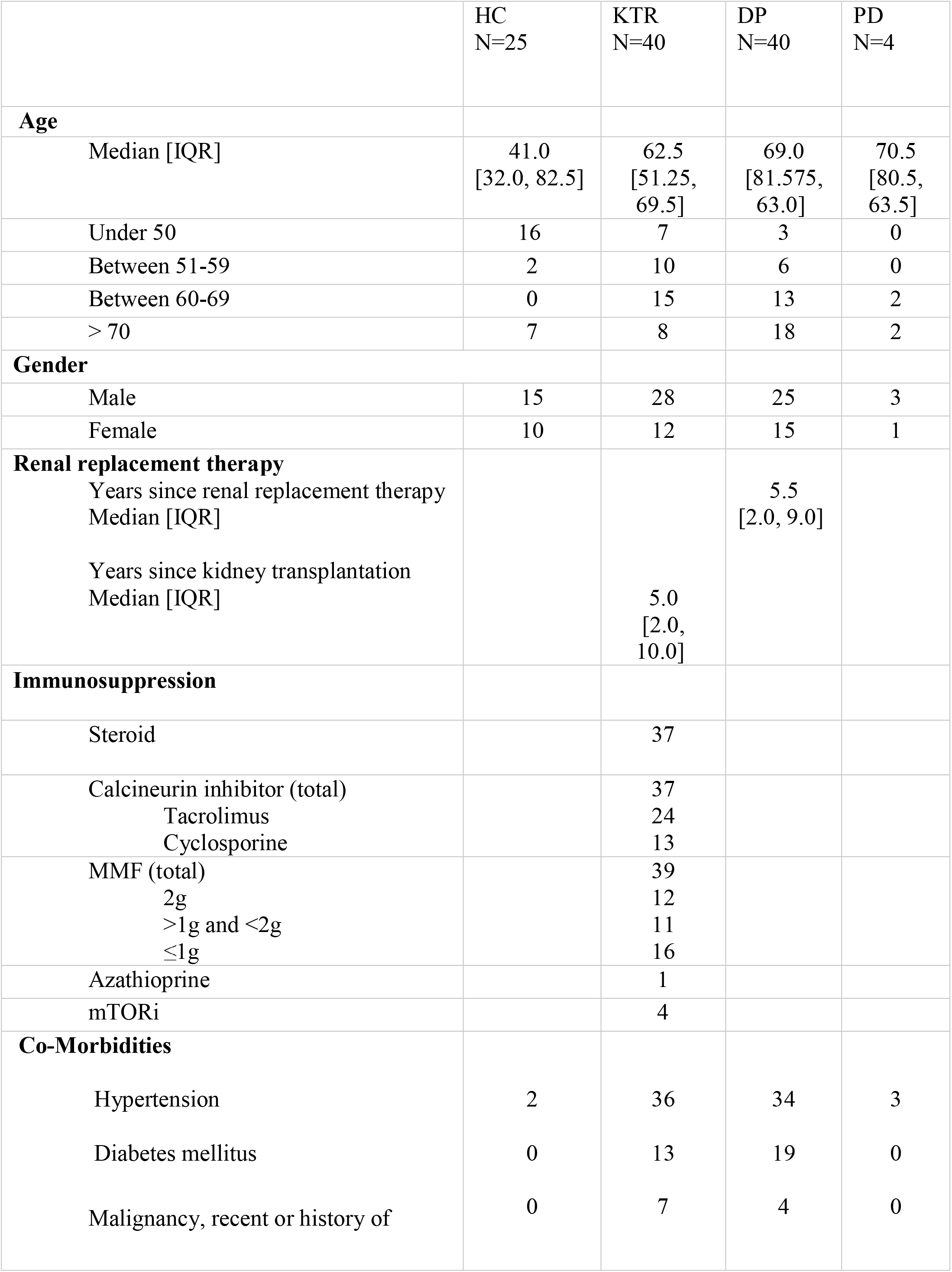
Patient characteristics

### DP and KTR show reduced B cells numbers but similar distribution among memory subsets prior vaccination (baseline)

B cell lymphopenia has been described for DP (*15*) and KTR (*16*) and might affect proper humoral immune responses. To initially address the frequency, distribution, and phenotype of peripheral blood B cells in DP, KTR compared to HC, we analyzed the distribution of B cell subsets at baseline (pre-vaccination) and 7±2 days after boost (Figure 1A). Of interest, the frequency of CD19+ B cells was significantly diminished only in KTR compared to DP at the assessment 7±2 days after boost, while otherwise no differences were observed (Figure 1B). However, substantial reductions of the absolute B cell counts were identified between KTR patients and HC at baseline as well as HC versus the DP and KTR cohorts, 7±2 days after boost, respectively (Figure 1C).

**Figure 1:**
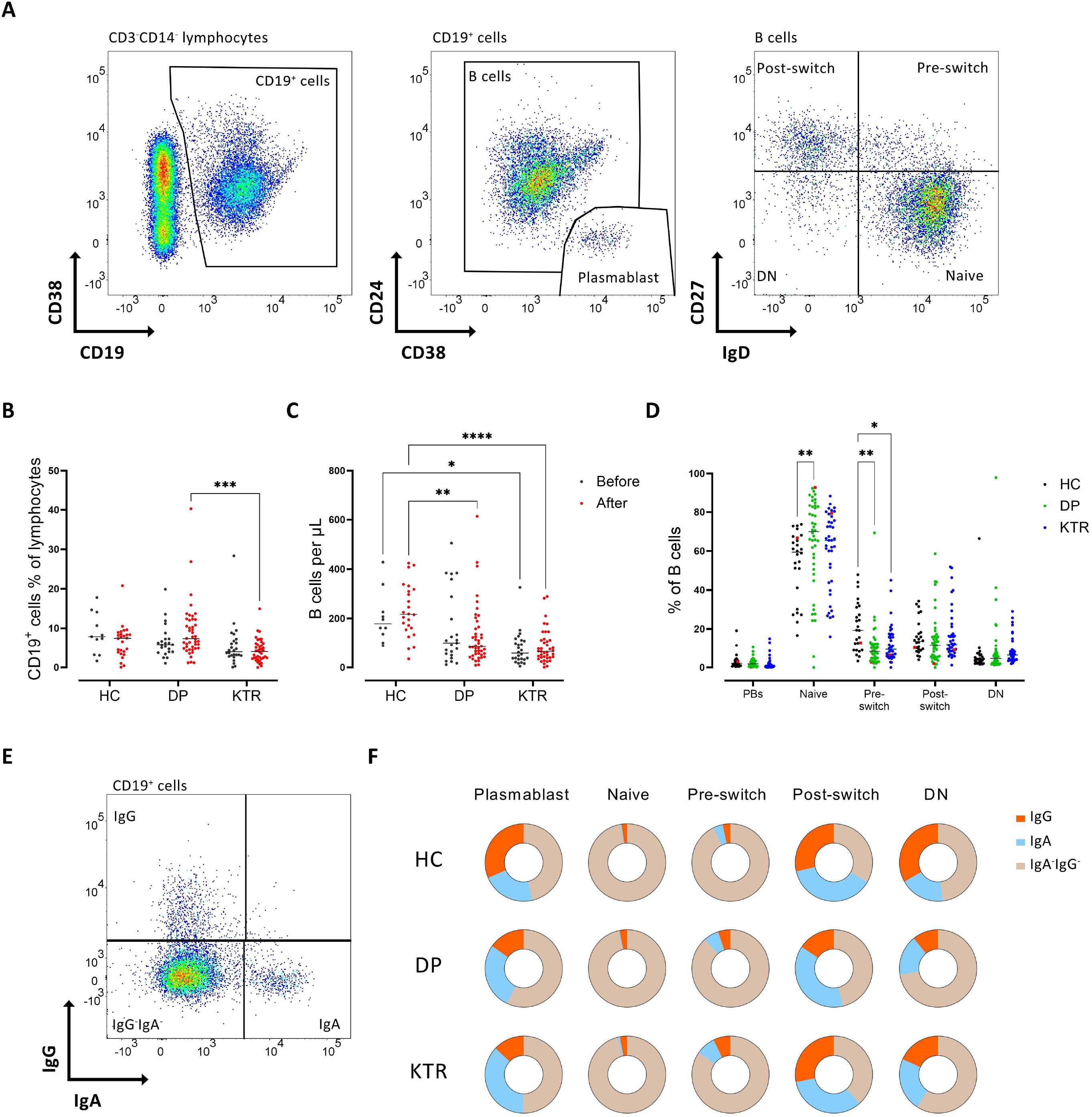
B cells are reduced in DP and KTR but show a similar distribution after BNT162b2 vaccination. Representative pseudocolor plots of CD19+ B cell gating into plasmablasts and mature B cells, and representative pseudocolor plots of IgD/CD27 based classification **(A)**. Frequency of CD19+ B cells (gates shown in (A)) in HC, DP and KTR **(B)** before vaccination and 7±2 days after 2nd vaccination with BNT162b2. Corresponding absolute numbers (per µl blood) measured by BD Trucount are shown in **(C)**. Frequencies of plasmablasts and according to CD27/IgD (gates shown in (A)) in mature B cells **(D)** 7±2 days after 2nd vaccination. Gating of immunoglobulin isotype expression of mature B cells from a HC. Previously infected individuals are indicated in red. **(E)**. Distribution of surface immunoglobulin isotype expression among HC, DP and KTR 7±2 days after 2nd vaccination **(F)**. Two-way ANOVA with Šidák’
ss post-test. *p<0.05, **p<0.01, ***p<0.001, ****p<0.0001.

The frequency of plasmablasts among total CD19+ B cells did not differ between groups (Figure 1D). DP and KTR patients carried lower frequencies of pre-switch B cells, while DP patients had an increased frequency of naïve B cells. Post-switch memory B cells as well as double negative (DN, CD27-IgD-) B cells did not differ significantly among groups (Figure 1D). Immunoglobulin isotype distribution in B cell subsets was not different among study groups (Figure 1E, F). In summary, KTR and DP showed a characteristic reduction of absolute B cells with certain differences in the pre-memory (naive and pre-switch) but no differences within B memory compartments.

### Substantially impaired serological response upon mRNA vaccination with BNT162b2 in DP and even more pronounced in KTR patients

Antibody response to BNT162b2 was assessed in all individuals 7±2 days after boost using the Euroimmun ELISA for the detection of IgG and IgA against the S1 domain of the SARS-CoV-2 spike. All HC seroconverted, were positive for both anti-S1 IgG and anti-S1 IgA (Figure 2 A, B), and showed SARS-CoV-2 neutralization (Figure 2C). Noteworthy, anti-S1 IgA and anti-S1 IgG titers were markedly diminished 7±2 days after boost in DP patients compared to HC (Figure 2A, B). In the S1 IgG assay, 31/44 (70.5%) of the DP were positive and 30/44 (68.2%) developed anti-S1 IgA antibodies.

**Figure 2:**
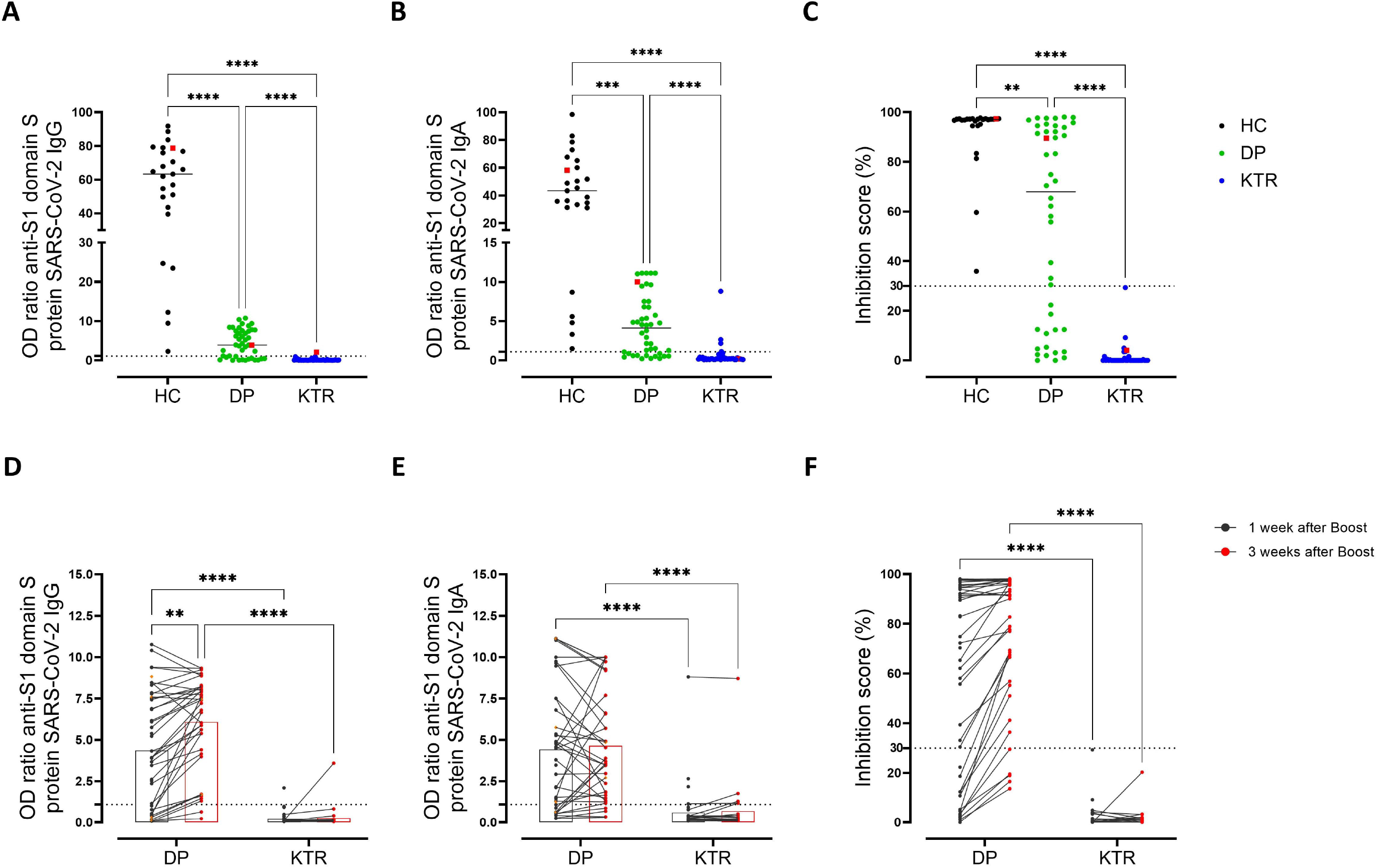
Humoral immune response is delayed in DP and markedly reduced in KTR. Humoral immune response against SARS-CoV-2 was assessed by Euroimmune ELISA for spike protein S1 IgG **(A)**, spike protein S1 IgA **(B)** and virus neutralization by a blocking ELISA **(C)** in HC (n=25), DP (n=44) and KTR (n=40) 7±2 days after 2nd vaccination with BNT162b2. Threshold of upper limit of normal is indicated as dotted lines. Follow-up sera were collected for 37 DP and 26 KTR patients, respectively 3-4 weeks after 2nd vaccination and investigated for S1 IgG **(D)**, IgA a **(E)** and virus neutralization **(F)**. (A-C) Kruskal-Wallis with Dunn’
ss post-test. Previously infected individuals are indicated in red. (D-F) Two-way ANOVA with Šidák’s post-test. *p<0.05, **p<0.01, ***p<0.001, ****p<0.0001.

Of particular interest, anti-S1 IgG and anti-S1 IgA responses were substantially diminished in KTR compared to HC and DP, respectively. Only one out of 40 patients (2.5%) was positive for IgG (apparently after prior unrecognized infection) and 4 patients for IgA (10%). Virus neutralization was observed in 30/44 (68.2%) DP patients (Figure 2C), while 0/40 KTR had inhibiting antiviral antibodies (Figure 2C). Interestingly, also the patient’s serum with IgG and prior infection did not achieve neutralizing effects. Previously infected individuals are indicated in red in Figure 2 (A, B, C). Their levels of antibody and neutralization is in the range of other individuals of the respective group.

In a previous report (*17*), HC showed no significant further increase of humoral response later than 28 days post initial vaccination with BNT162b2. A delayed immune response might have explained the initial low seroresponse in immunocompromised individuals (DP and KTR) with mRNA vaccines. Therefore, we collected a further follow-up for KTR and DP 3-4 weeks after boost. Interestingly, anti-S1 IgG and surrogate neutralization increased significantly in DP (Figure 2D), while anti-S1 IgA remained stable (Figure 2E) during this additional observation time. In contrast, KTR patients did not develop additional anti-S1 IgG, anti-S1 IgA antibodies and neutralization capacity until the second follow-up investigation 3-4 weeks after the boost (Figure 2D-F). In summary, KTR showed a significantly reduced serological response including lack of further increases up to 3-4 weeks after BNT162b2 boost.

### Impaired induction of anti-BNT162b2 B cell and plasma cell responses in KTR and HD patients

In order to better understand the underlying B and plasma cell differentiation upon vaccine challenge, we developed a flow cytometric method to identify and quantify RBD-specific B cells in human peripheral blood. B cells (CD3−CD14−CD19+) able to bind simultaneously RBD-AF488 and RBD-AF647 have been validated as antigen-specific (Figure 3A). The specificity of RBD binding was further confirmed by blocking with unlabeled RBD prior to staining (Figure 3A). Subsequently RBD+ B cells were further analyzed according to their distribution among subsets and isotypes (gated as shown for general B cells in Figure 1A, E).

**Figure 3:**
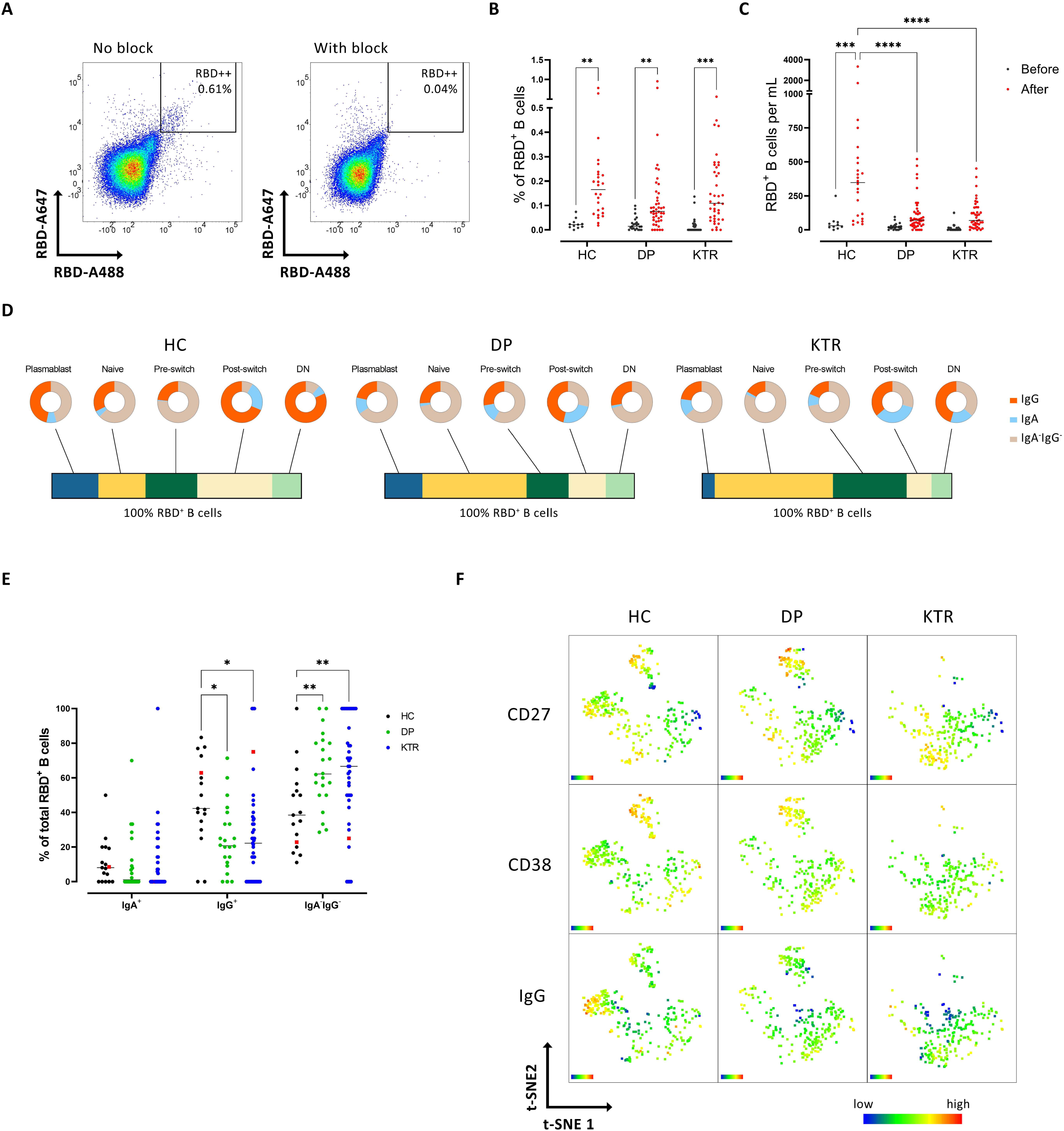
RBD-specific B cells are present in DP and KTR patients after BNT162b2 vaccination but populate different B cell subsets. Representative dot plot of double positive cells RBD-specific B cells before and after blocking with unlabeled RBD are shown **(A)**. The frequencies **(B)** and absolute numbers **(C)** of RBD+ cells among total CD19+ B cells measured before and 7±2 days after 2nd vaccination. Frequencies of plasmablasts, naïve, pre-switch, post-switch and double negative B cells (bar) and immunoglobulin isotype distribution among subsets (cakes) **(D)**. Immunoglobulin isotype expression among total RBD+ cells in HC, DP and KTR and 7±2 days after 2nd vaccination **(E)**. Two dimensional t-SNE of all RBD+ cells in HC (n=21), DP (n=23) and KTR (n=34). Color code indicates expression of CD27 (upper panel) CD38 (middle panel) and IgG (lower panel) **(F)**. Previously infected individuals are marked red (E). (B-D) Two-way ANOVA with Šidák’
ss post-test. (E) Kruskal-Wallis with Dunn’s post-test. *p<0.05, **p<0.01, ***p<0.001, ****p<0.0001.

Overall, an increased frequency of RBD-specific B cells among CD19+ B cells was found 7±2 days after boost compared to baseline for HC, DP and KTR (Figure 3B). The absolute number of antigen-specific B cells was significantly increased only in HC at 7±2 days after boost in contrast to DP and KTR patients (Figure 3C).

Subsequent analyses addressed the distribution of the RBD-specific B cells among B cell subsets (gating as seen in Figure 1A). Most notably, a large number of RBD+ B cells was found in the plasmablast compartment in HC, which was significantly lower in DP and KTR (Figure 3D and Figure S2A). The very limited antigen-specific B cells in KTR resided preferentially within the naïve and pre-switch compartment compared to HC (Figure 3D and Figure S2A). In contrast, antigen-specific B cells from HC were detected mainly within post-switch and among double negative B cells and belonging largely to the memory compartment (Figure 3D). Consistent with impaired (not completely executed) B memory induction, the frequency of IgM RBD+ B cells (defined as IgG-IgA-) was more often detected in KTR and DP patients compared to HC, in whom antigen-specific IgG+ B cells dominated. The frequency of IgA+ RBD+ B cells was comparable across groups (Figure 3E).

Two-dimension t-SNE plots clustering all RBD+ B cells according to expression patterns, analyzed with a color axis for CD27, CD38 and IgG, illustrate the notable differences between groups including the substantially reduced plasmablasts (CD27++, CD38++) and IgG expressing RBD+ B cells in the KTR cohort (Figure 3F). In summary, KTR patients were not only characterized by a reduced overall number of antigen specific B cells, but also exhibited signatures of inappropriate B cell memory induction.

### Unique correlation of anti-BNT162b2 serological and B cell responses

Our earlier vaccination studies against tetanus, diphtheria and KLH (Keyhole Limpet Hemocyanin) did not reveal a typical relation between plasmablast/ B cell response and the serologic Ig outcome (*6-8*) in contrast to such relation for polysaccharides, such as meningococcal and pneumococcal vaccine (*18, 19*). Therefore, we wondered how the anti-BNT162b2 humoral immune and B cell specific responses against an mRNA vaccine are interrelated. Therefore, a correlation matrix including all groups and patients was carried out. As previously described (*20*), the neutralization capacity strongly correlated with anti-S1 IgG as well as anti-S1 IgA (Figure 4A). Interestingly, the frequency of total RBD+ and absolute number of RBD+ cells did not correlate with anti-S1 IgG, IgA and neutralization capacity, respectively. However, and most noteworthy, both, the frequency and total number of RBD+ plasmablasts correlated with all parameters of humoral response (anti-S1 IgG, anti-S1 IgA and the neutralization capacity). Subsequent analyses addressed how non-responders with a negative neutralization test (<30%) differ from responders (>30%). Responders and non-responders were significantly different in the frequency and number of RBD+ plasmablasts and RBD+ pre-switch memory B cells as well as in the frequency of RBD+ naïve B cells (Figure 4B). This data suggests clear interdependence of the distinct memory B and plasma cell compartments characteristic for this mRNA vaccine.

**Figure 4.**
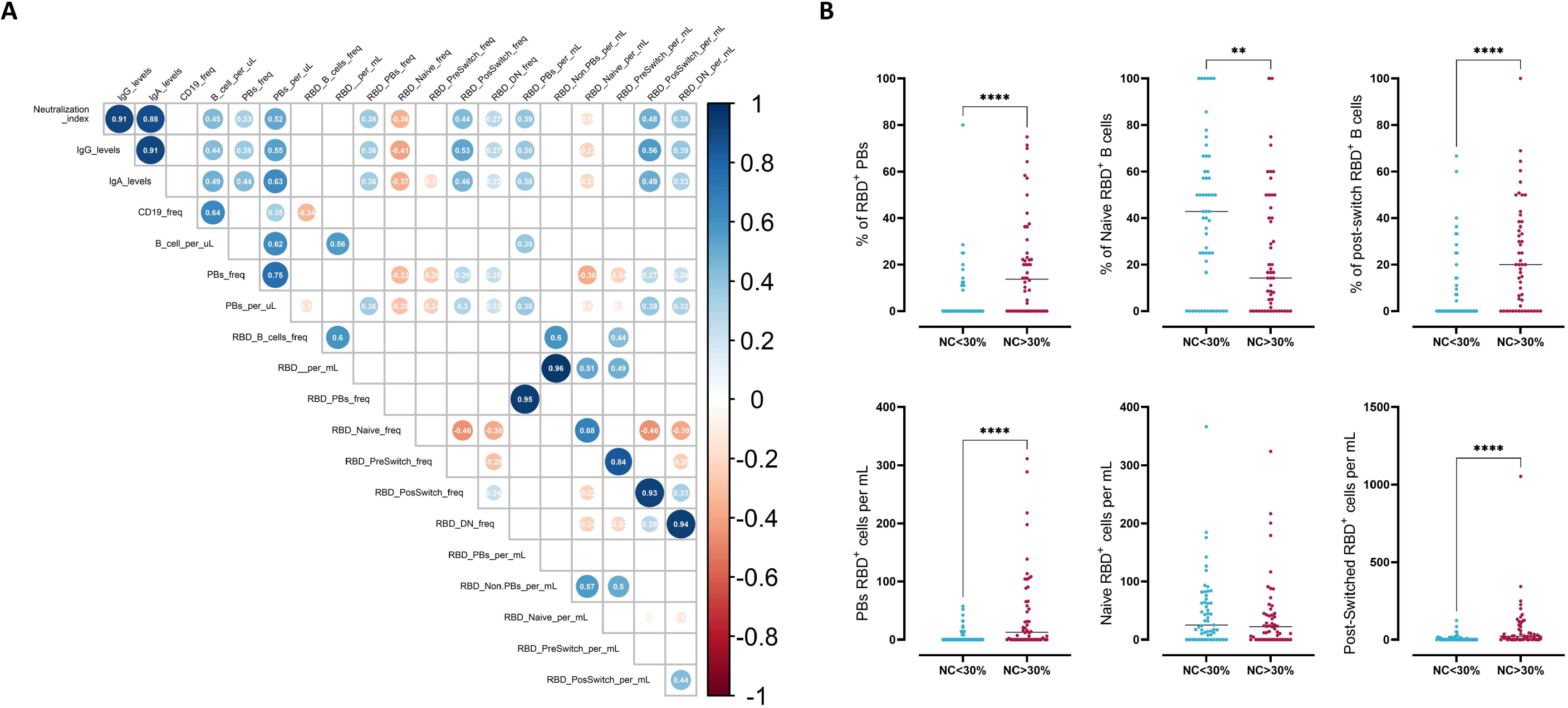
Correlation of anti-BNT162b2 serological and B cell responses. Spearman’
ss correlation matrix showing the correlation of frequency of RBD+ cells in each B cell subset in the cohort. Corresponding correlations are represented by red (negative) or blue (positive) circles; size and intensity of color refer to the strength of correlation **(A)**. Only correlations with p ≤ 0.05 are indicated. Frequency (upper panel) and absolute numbers (lower panel) of RBD+ plasmablasts (PB), naïve B cells and post-switch B cells in non-responders (surrogate virus neutralization capacity (NC) NC<30) and responders NC (>30%) **(B)**. Each point represents a donor. Unpaired two-sided Mann-Whitney U test. *p<0.05, **p<0.01, ***p<0.001, ****p<0.0001.

### Diminished T cell and plasmablast response in KTR

To further understand the lack of B cell memory induction, we sorted CD27++CD38++ plasmablasts, CD27+CD38var memory B cells and HLA-DR+CD38+ activated T cells of the peripheral blood as indicators of the ongoing immune response after vaccination (*7, 21*) and generated single cell transcriptomes combined with Cellular Indexing of Transcriptomes and Epitopes by Sequencing (CITE-seq) (*22*) for selected surface markers (Figure S3) 7±2 days after boost vaccination. After removal of doublets, we analyzed a total of 10796 cells. According to their transcriptomes, these cells were categorized in 10 different clusters as shown by Uniform Manifold Approximation and Projection for Dimension Reduction (UMAP) (*23*) (Figure 5A). We focused on the most abundant clusters. Four of which belong to the T cell compartment, with the most abundant cluster 0 representing activated *CD8*+*HLADR*+*MS4A1*+ T cells, followed by cluster 1 representing different types of the CD4+ T cells, including e.g. *FOXP3*+*CD25*+ regulatory T cells (Figure 5C, Figure S3). Cluster 2 contains different populations of CD8+ T cell expressing either CD45RA or CD45RO (Figure S3) and finally cluster 5 representing a *TCF7*+*CD27*+*GZMK*+ T cells. Cluster 6 contains proliferating cells expressing *MKI67* (Figure 5B-C). Memory B cells expressing high levels of *MS4A1, HLA-DRA* and *CD27* are located in cluster 3. Cluster 4 represents plasmablasts expressing *CD27, CD38, PRDM1* and *IRF4* (Figure 5B-C). Of note, the protein expression of selected surface markers detected by CITE-seq support the classification of the 7 main clusters (Figure S3). In the selected cohort of 4 KTR, 3 did not show a serological response to the vaccination (non-responder), while one individual had detectable anti-S1 IgG antibodies but also a previously undetected infection (responder). Accordingly, the 3 non-responders had reduced frequencies of cluster 4, 5 and 6 representing plasmablasts, *TCF7*+*CD27*+*GZMK*+ T cells and proliferating *MKI67*-expressing lymphocytes (Figure 5D-E). CD45RO+ follicular T helper like cells expressing either *IL21* or *PDCD1*+ could only be detected in the CD4+ T cell compartment (Cluster 1) of the non-responder #3 and the responder (Figure 5F). Interestingly, the majority of memory B cells (cluster 3) of the responder expressed *ITGAX* (gene encoding for CD11c), while this subpopulation in cluster 3 was almost absent in the non-responders.

**Figure 5:**
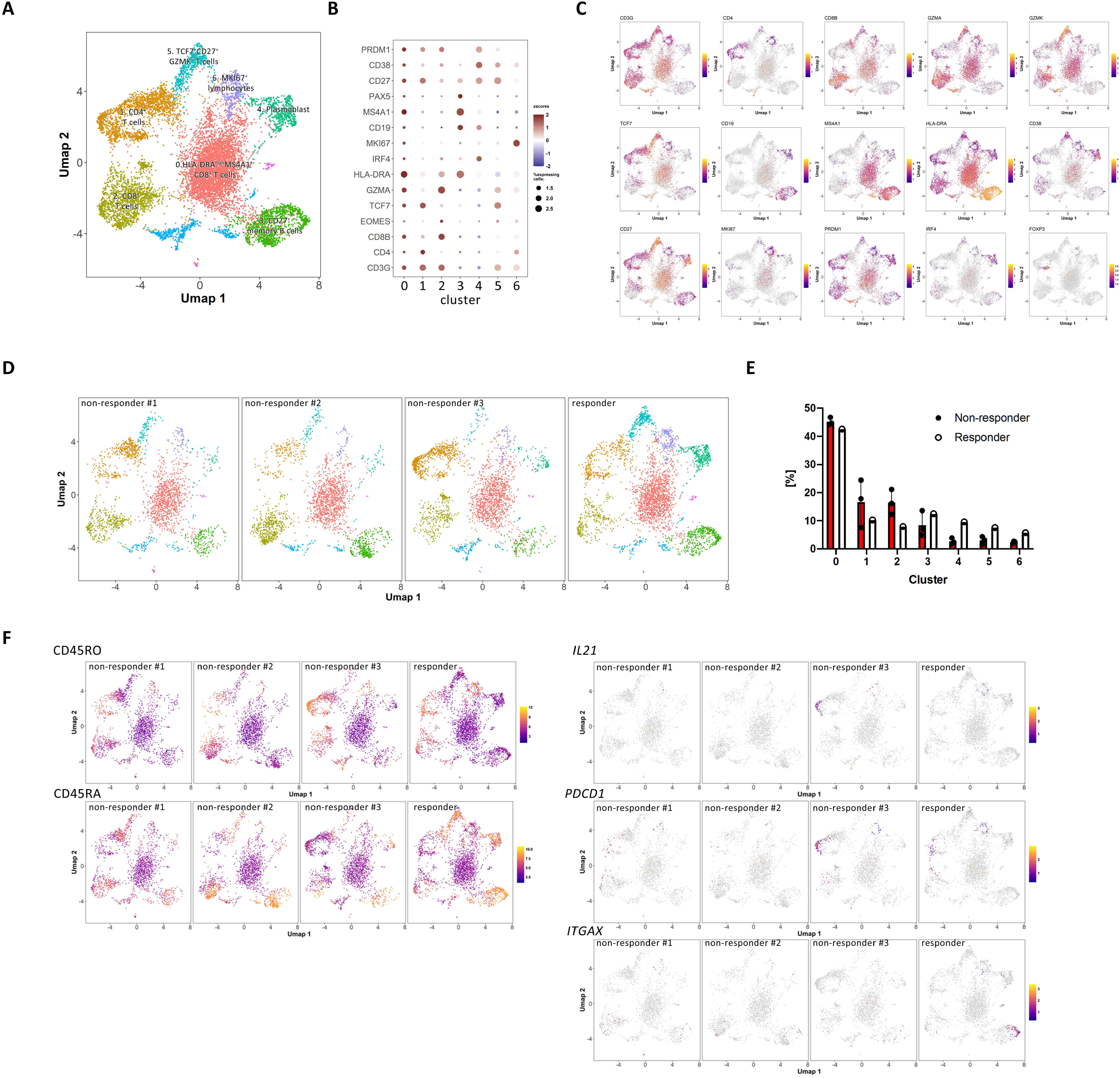
Diminished T cell and plasmablast response in KTR. UMAP clustering of 10796 CD27++CD38++ plasmablasts, CD27+CD38var memory B cells and HLA-DR+CD38+ activated T cells from 3 non-responders and 1 responder (KTR) 7±2 days after boost vaccination **(A)**. Relative expression levels of selected signature genes in the 7 identified clusters **(B)**. UMAP representation of the expression levels of selected signature genes for pooled cells **(C)**. UMAP clustering from 3 non-responder and 1 responder (KTR) **(D)**. Cluster frequency comparison between the 3 non-responder and the 1 responder KTR **(E)**. UMAP representation of the expression levels of CD45RO and CD45RA proteins and *IL21, PDCD1* and *ITGAX* genes in the 3 non-responder and 1 responder (KTR) **(F)**

## DISCUSSION

SARS-CoV-2 mRNA vaccines are proven highly effective in clinical trials for protection against COVID-19 (*2, 24*), while it is not yet clear how these vaccines induce and maintain B cell memory responses in normal controls as well as among immunocompromised patients. Available data showed a durable humoral response (*25*) in healthy individuals and also elderly patients but largely limited to Ig titers and T cell response (*2, 17*). Moreover, mRNA vaccines have been reported to induce germinal center (GC) responses in mice and are expected to result in lasting plasma cell responses (*26*). Here we present a first study investigating the distribution of anti-BNT162b2 antigen-specific B cell responses among HC in comparison to DP and KTR as prototypes for differentially immunocompromised patients. An assay for the detection of RBD-specific B lineage cells has been developed with high specificity based on our prior experiences for the detection of antigen-specific B cells and plasmablasts (i.e. tetanus, KLH, pentraxin (*8, 27*)). Of particular interest, seroconversion and the induction of neutralizing antibodies upon BNT1622b vaccination was very robust and similar in our cohort in HC as observed in prior studies (*2*). In these individuals we also found a typical induction of antigen-specific plasmablasts and post-switch memory B cells upon vaccination boost. Thus, the findings among HC provide a valid comparison with the two patient cohorts. Among those, we observed a markedly diminished generation of antigen-specific B lineage cells in KTR and DP patients especially within major effector compartments of protective B cell immunity, namely plasmablasts and memory B cells. Consistently, lower IgG+ anti-RBD cells were found related to impaired induction of a vaccine response. This was accompanied by a low rate of seroconversion of DP on day 7±2 that somewhat improved 3-4 weeks after vaccination. In the KTR cohort, the rate of serological and cellular response was almost absent with only one patient developing specific anti-S1 IgG but apparently based on a prior unrecognized infection and likely reflecting a further boost. Interestingly, no KTR patient exhibited a positive neutralization at day 7± 2 after boost including the patient with positive IgG titers. Also, at a further follow up 3-4 weeks after boost no seroconversions have been found in KTR. These data are in contrast to recently published data of Boyarsky et al. who described a seroconversion in 23/223 (10.35%) of KTR vaccinated with BNT162b2 already after the first dose (*5*). For DP patients, seroconversion rates of about 90% after two doses have been described which is similar to the findings in our cohort (*4, 28*). The ongoing antigen exposure through mRNA vaccines allows a prolonged GC maturation and a second wave of GC response (*26*), which might be an explanation for the further increase in antibody titers in DP. In KTR this ongoing maturation is lacking. The large discrepancy of the serological response and impaired B cell response in our KTR cohort raises the question, which factors or mechanism are involved. Incorrect handling of the sensitive mRNA vaccine can be excluded since the same mRNA vaccine of the same lot mounted a favorable response among DP patients. The major difference between KTR and DP/HC appeared to be the almost uniform immunosuppressive therapy with MMF, CNI and glucocorticoids in the KTR patients, although the individual impact of MMF, cyclosporine vs. tacrolimus and glucocorticoids needs to be further delineated. However, these drugs impact on T and B cells, respectively, including proper induction of plasma cells. In this regard, single cell transcriptomes and CITE-seq analyses identified substantial differences between the vaccine responder and three non-responders within the KTR cohort. Namely plasmablasts, *TCF7*+*CD27*+*GZMK*+ T cells and proliferating *MKI67*-expressing lymphocytes were increased in the responder suggesting that these three subsets are key drivers of a successful BNT162b2 response.

Three individuals with likely prior asymptomatic virus exposure (each in HC, DP and KTR cohorts) were identified by preexisting antibodies against viral NC. Interestingly, they did not show notable differences in vaccine response compared to their cohorts, with the exception of one seroconversion in the KTR. A number of recent studies have reported that a first vaccination in previously infected individuals acts as a boost that mounts antibody levels over individuals vaccinated twice with an mRNA vaccine. It further does not change isotype distribution among memory B cells (*29-31*). After a natural COVID-19 infection the numbers of RBD-specific memory B cells of healthy individuals are similar to those of patients who recovered from natural COVID-19 infection, while levels of anti-S1 and anti-RBD IgG are significantly higher in vaccinated individuals(*32*). Interestingly, the clonality of IGLV genes among RBD+ was very comparable between natural infection and the mRNA vaccination (*32*). At the moment it is not known whether the magnitude of antibody levels or the presence or magnitude of a T cell response, or both, correlate with the protection against symptomatic COVID-19. However, after natural COVID-19 infection the presence of anti-spike antibodies protects from recurrent infection, while a previous infection without detectable antibodies does not (*33*). Initial observational data from Israel indicate that CKD as well as immunosuppression have a negative impact on vaccine efficacy (*34*), which indicates that the lack of B cell memory also results into diminished protection.

Interestingly, we find a unique relation between plasmablast response and Ig titers which is not seen in tetanus and KLH vaccination (*6-8*) and has only been described for polysaccharide and protein-polysaccharide conjugate vaccinations (*35*). Polysaccharide vaccines in contrast to mRNA vaccines mounted certain serological responses following vaccination in KTR (*36*) which possibly relates to T independent responses. In this context, our single cell analysis confirmed a diminished plasmablast induction in the non-responders (KTR) but importantly also lack of proper CD4 and CD8 T cell activation. In the responder, classical signs of vaccine response with simultaneous B and T cell activation occurred but insufficient to induce neutralizing antibody titers. KTR have to be considered as a patient group who remains vulnerable to SARS-CoV-2 infection, even if vaccinated with the currently established BNT162b2 vaccination scheme.

In summary, we describe a diminished humoral response to BNT162b2 as well as lack of appropriate memory formation including RBD-specific plasmablasts and post-switch memory B cells in DP and KTR. DP are able to further mature their antibody response in contrast to KTR. Our data provide evidence that proper activation of *TCF7*+*CD27*+*GZMK*+ T cells and *MKI67*-expressing lymphocytes are required for the induction of B and plasma cell memory upon the mRNA vaccine. As such, optimized vaccination strategies especially for KTR patients are needed to achieve adequate antiviral protection.

## MATERIALS AND METHODS

### Study design

The study was designed to investigate seroresponse (anti-S1 IgA, anti-S1 IgG levels and neutralization test) and B cell memory formation in DP and KTR after vaccination with BNT162b2. Blood samples were obtained before vaccination as well 7 ± 2 days and 3-4 weeks after the second dose.

### Study participants

Peripheral blood samples (EDTA anti-coagulated or serum-tubes, BD Vacutainersystem, BD Diagnostics, Franklin Lakes, NJ, USA) from 25 healthy controls, 40 hemodialysis patients, 4 peritoneal dialysis patients and 40 kidney transplant recipients were collected at 7 ± 2 days after the second dose of SARS-CoV-2 BNT162b2 vaccination. Individuals of KTR and DP are in part also represented in published manuscripts (serological response only) (*37, 38*). Material before vaccination was available for 19 healthy controls, 21 hemodialysis patients, 2 peritoneal dialysis patients, and 28 kidney transplant recipients, while a follow up after 3-4 weeks after boost is available for currently for 26 KTR and 37 DP. Donor information is summarized in Table 1. All participants gave written informed consent according to the approval of the ethics committee at the Charité University Hospital Berlin (EA2/010/21, EA4/188/20), the ethics committee of Saxony-Anhalt (EA7/21) and the ethics committee of the University of Greifswald (BB019/21).

### Samples processing and isolation of peripheral blood mononuclear cells (PBMCs) and staining

Serum tubes were centrifuged at 3000rpm for 10 minutes to separate plasma. Serum was stored at -20C for antibody analysis. PBMCs were prepared by density gradient centrifugation using Ficoll-Paque PLUS (GE Healthcare Bio-Sciences, Chicago, IL, USA). For staining 1-3 x 10^6^ cells were suspended in 50□µl of PBS/0.5% BSA/EDTA and 10□µl Brilliant Buffer (BD Horizon, San Jose, CA, USA. Cells were stained for 15_L_min on ice and washed afterwards with PBS Dulbecco containing 1% FCS (fetal calf serum, Biowest, Nuaillé, France) (810 xg, 8□min, 4°C). Flow cytometric analysis was performed as indicated in the Figures 1-4.

### Analytical methods

All flow cytometry analyses were performed using a BD FACS Fortessa (BD Biosciences, Franklin Lakes, NJ, USA). To ensure comparable mean fluorescence intensities (MFIs) over time of the analyses, Cytometer Setup and Tracking beads (CST beads, BD Biosciences, Franklin Lakes, NJ, USA) and Rainbow Calibration Particles (BD Biosciences, Franklin Lakes, NJ, USA) were used. For flow cytometric analysis, the following fluorochrome-labeled antibodies were used: BUV737 anti-CD11c (BD, clone B-ly6, 1:50), BUV395 anti-CD14 (BD, clone M5E2, 1:50), BUV395 anti-CD3 (BD, clone UCHT1, 1:50), BV786 anti-CD27 (BD, clone L128, 1:50), BV711 anti-CD19 (BD, clone SJ25C1,1:25), BV605 anti-CD24 (BD, clone ML5, 1:50), BV510 anti-CD10 (BD, clone HI10A, 1:20), BV421 anti-CXCR5 (BD, clone RF8B2, 1:20), PE-Cy7 anti-CD95 (ThermoFischer, Waltham, MA, USA clone APO-1/Fas, 1:100), PE-CF594 anti-IgD (Biolegend, San Diego, CA, USA, clone IA6-2, 1:5000), APC-Cy7 anti-CD38 (Biolegend, clone HIT2, 1:1000), PE-Cy7 anti-IgG (BD, clone G18-145, 1:1000), anti-IgA-Biotin (BD, clone G20-359, 1:50), BV650 anti-IgM (BD, clone MHM-88, 1:50), FITC anti-HLA-DR (Biolegend, clone L234, 1:25), PE anti-CD21 (BD, clone B-ly4, 1:25), APC anti-CD22 (BD, clone S-HCL-1, 1:25). Siglec-1 (CD169, 1:25) expression analysis on CD14+ monocytes was performed at baseline and at the follow-up time-point as previously described (11). Number of absolute B cells was measured with Trucount (BD) and samples were processed according to the manufacturer’s instruction.

### Staining of antigen-specific B cells

To identify RBD-specific B cells, recombinant purified RBD (DAGC149, Creative Diagnostics, New York, USA) was labeled with either AF647 or AF488. Double positive cells were considered as antigen-specific (Figure 3). Antigens were labeled at the German Rheumatism Research Centre (DRFZ), Berlin with NHS esters conjugation for AF647 and AF488. A blocking experiment using unlabeled RBD in 100-fold concentration was used to ensure specificity of the staining (Figure 3A).

### Enzyme-linked immunosorbent assay (Euroimmun)

The Euroimmun anti-SARS-CoV-2 assay is a classical enzyme-linked immunosorbent assay (ELISA) for the detection of IgG to the S1 domain of the SARS-CoV-2 spike (S) protein, IgA to the S1 domain of the SARS-CoV-2 spike protein, and IgG to the SARS-CoV-2 NCP protein. The assay was performed according to the manufacturer’
ss instructions and as described previously (*20*). Briefly, serum or plasma samples were diluted at 1:100 in sample buffer and pipetted onto strips of 8 single wells of a 96-well microtiter plate, precoated with recombinant SARS-CoV-2 spike or nucleocapsid proteins. A calibrator, a positive control and a negative control were carried out on each plate. After incubation for 60 minutes at 37°C, wells were washed 3 times and the peroxidase-labelled anti-IgG or anti-IgA antibody solution was added, followed by a second incubation step for 30 min. After three additional washing steps, substrate solution was added and the samples incubated for 15 - 30 minutes in the dark. After adding the stop solution, optical density (OD) values were measured on a POLARstar Omega plate reader (BMG Labtech, Ortenberg, Germany) at 450 nm and at 620 nm. Finally, OD ratios were calculated based on the sample and calibrator OD values. For all analytes, a ratio < 0.8 was considered to be non-reactive or negative. An OD-ratio of ≥ 1.1 was considered to be positive for all three analytes. Dilutions of 1:10 were prepared when ELISA when values were close to the saturation point of the respective ELISA.

### Surrogate SARS-CoV-2 neutralization test (GenScript)

This blocking ELISA qualitatively detects anti-SARS-CoV-2 antibodies suppressing the interaction between the receptor binding domain (RBD) of the viral spike glycoprotein (S) and the angiotensin-converting enzyme 2 (ACE2) protein on the surface of cells. After pre-incubation of samples and controls, which allows antibodies in the serum to bind to a horseradish peroxidase (HRP)-conjugated RBD fragment (HRP-RBD), the mixture is added to a capture plate coated with human ACE2 protein. Any unbound HRP-RBD or HRP-RBD bound to non-neutralizing antibodies is captured on the plate. Complexes of neutralizing antibodies and HRP-RBD do not bind on the plate and are removed after three washing steps. Then, TMB is added as a substrate, allowing HRP to catalyze a colour reaction. The colour of the solution changes from blue to yellow after addition of the stop reagent, and can be read by a mictotiter plate reader at 450nm (OD450). The absorbance of the sample is inversely correlated with the amount of SARS-CoV-2 neutralizing antibodies. Positive and negative controls serve as internal controls, the test is considered valid only if the OD450 for each control falls within the respective range (OD450negative control > 1.0, OD450positive control < 0.3). For final interpretation, the inhibition rates were determined using the following formula: Inhibition score (%) = (1 - (OD valuesample/OD valuenegative control) x 100%). Unless stated otherwise, scores < 30% were considered negative, scores ≥ 30% were considered positive.

### Sorting of plasmablast, B and T cells from peripheral blood for single cell analysis

Cells were enriched from peripheral blood using StraightFrom Whole Blood CD19, CD3 and CD138 MicroBeads (Miltenyi Biotec) according to manufacturer’s instructions. Enriched cells were incubated with Fc Blocking Reagent (Milteniy Biotec) following manufacturer’s instructions and subsequently stained up to 10^7^ lymphocytes per 100µL for 30min at 4°C with the following fluorophore-conjugated anti-human antibodies:, VioBlue anti-CD14 (Miltenyi Biotec, Clone TÜK4, 1:200), VioBlue anti-CD16 (Miltenyi Biotec, clone VEP13, 1:50), FITC anti-CD3 (DRFZ, clone UCHT1, 1:100), PE anti-CD27 (Miltenyi Biotec, clone MT271, 1:50), APC anti-CD38 (BioLegend, clone HIT2, 1:25), and the following TotalSeq-C oligomer-conjugated anti-human antibodies: CD138 (clone DL-1021, 1:50), CD20 (clone 2H7, 1:1000), CD4 (clone SK3, 1:100), CD8 (clone SK1, 1:1000), CD45RO (clone UCHL1, 1:100), CD45RA (clone HI100, 1:400), CD25 (clone BC96, 1:100), CD127 (clone A019D5, 1:400) and CD154 (clone 24-31, 1:100), all from BioLegend. To allow for sequencing of pools of cells from different donors, cells were also incubated with TotalSeq oligomer-conjugated hashtag antibodies (TotalSeq-C anti-human Hashtag antibody 1 to 4 from BioLegend, 1:250). DAPI was added before sorting to allow dead cell exclusion. All sorting were preformed using a MA900 Multi-Application Cell Sorter (Sony Biotechnology). Cell counting was performed using a MACSQuant flow cytometer (Miltenyi Biotec). Sorted populations were identified as plasmablasts (DAPI-CD3-CD14-CD16-CD38++CD27++), memory B cells (DAPI-CD3-CD14-CD16-CD38varCD27+) and activated T cells (DAPI-CD3+CD14-CD16-CD38+HLA-DR+). The three sorted populations were pooled in equal proportions and further processed for single cell RNA sequencing.

### Single Cell RNA-library preparation and sequencing

Single cell suspensions were obtained by cell sorting and applied to the 10x Genomics workflow for cell capturing and scRNA gene expression (GEX) and CiteSeq library preparation using the Chromium Single Cell 5’ Library & Gel Bead Kit as well as the Single Cell 5’ Feature Barcode Library Kit (10x Genomics). After cDNA amplification the CiteSeq libraries were prepared separately using the Single Index Kit N Set A. Final GEX libraries were obtained after fragmentation, adapter ligation and final Index PCR using the Single Index Kit T Set A. Qubit HS DNA assay kit (life technologies) was used for library quantification and fragment sizes were determined using the Fragment Analyzer with the HS NGS Fragment Kit (1-6000bp) (Agilent). Sequencing was performed on a NextSeq500 device (Illumina) using High Output v2 Kits (150 cycles) with the recommended sequencing conditions for 5’ GEX libraries (read1: 26nt, read2: 98nt, index1: 8nt, index2: n.a.) and Mid Output v2 Kits (300 cycles).

### Single-cell transcriptome Sequencing

Raw sequence reads were processed using cellranger-5.0.0, including the default detection of intact cells. The number of expected cells was set to 3000. Mkfastq and count were used in default parameter settings for demultiplexing and quantifying the gene expression. Refdata-cellranger-hg19-1.2.0 was used as reference. The cellranger output was further analyzed in R using the Seurat package (version 3.2.2) (*39*). In particular, transcriptome profiles for 4 KTR were merged, normalized, variable genes were detected and a Uniform Manifold Approximation and Projection (UMAP) was performed in default parameter settings using FindVariableGenes, RunPCA and RunUMAP with 30 principle components. Expression values are represented as ln (10,000 * UMIsGene) /UMIsTotal +1). Transcriptionally similar clusters were identified using shared nearest neighbor (SNN) modularity optimization with a SNN resolution of 0.1 resulting in x clusters. Annotation of clusters was performed manually by visual inspection of typical marker genes for cell-types and cell-states. Cells from different patients were identified according to their Hashtag expression, resulting in 2275, 3619, 2322, and 2580 cells for the Hashtags1-4 respectively. Data from transcriptome sequencing and immune profiling in GEO under the accession GSEXXXXX.

### Data Analysis and Statistics

All samples included in the final analyses had at least 1 × 10^6^ events with a minimum threshold for CD19+ cells of 5,000 events. Flow cytometric data were analyzed by FlowJo software 10.7.1 (TreeStar, Ashland, OR, USA). For the presentation of t-SNE plots, a concatenated file composed of all RBD+ cells from HC, DP, and KTR was used. GraphPad Prism Version 5 (GraphPad software, San Diego, CA, USA) was used for statistical analysis. CD20, CD24, CD27, CD38, CXCR5, IgD, IgA and IgG markers were used for t-SNE clustering analysis. For multiple group comparison, two-way ANOVA with Šidák’s post-test for multiple comparison or Kruskal-Wallis with Dunn’
ss post-test was used. Spearman correlation coefficient was calculated to detect possible associations between parameters or disease activity, respectively. P-values < 0.05 were considered significant. Correlation matrix was calculated using base R and corrplot package (R Foundation for Statistical Computing) using Spearman method.

### Supplementary Materials

Supplementary figures:

Figure S1: Nucleocapside among all donors.

Figure S2: Subset distribution of RBD+ B cells of HC, KTR and DP

Figure S3: Expression of surface markers in 4 KTR by Cellular Indexing of Transcriptomes and Epitopes by Sequencing (CITE-seq).

## Data Availability

All data are available in the main text or the supplementary materials.

## Acknowledgments

The authors are grateful to Dr. Michael Moesenthin, Dr. Peter Bartsch (both Dialysezentrum Burg), Dr. Ralf Kühn, Dr. Dennis Heutling (both Dialyse Tangermünde), Dr. Petra Pfand-Neumann (Nierenzentrum Köthen) and Dr. Jörg-Detlev Lippert (MVZ Diaverum, Neubrandenburg) for patient recruitment as well as Dr. Petra Glander and Pia Hambach for biobanking of samples.

## Funding

Federal Ministry of Education and Research (BMBF) grant BCOVIT, 01KI20161 (ES).

The Berlin Institute of Health with the Charité Clinician Scientist Program funded by the Charité –Universitätsmedizin Berlin and the Berlin Institute of Health (ES)

The Berlin Institute of Health with the Starting Grant-Multi-Omics Characterization of SARS-CoV-2 infection, Project 6 “Identifying immunological targets in Covid-19” (MFM).

Ministry for Science, Research and Arts of Baden-Württemberg, Germany (HS)

German Society of Rheumatology grant (ALS)

Sonnenfeldstiftung grant, Berlin, Germany (AS, KK)

Deutsche Forschungsgemeinschaft grants KO 2270/7-1, KO-2270/4-1 (KK); Do491/7-5, 10-2, 11-1, Transregio 130 TP24 (TD)

State of Berlin and the “European Regional Development Fund” with the grant ERDF 2014– 2020, EFRE 1.8/11, Deutsches Rheuma-Forschungszentrum (MFM)

The Deutsche Forschungsgemeinschaft (DFG) through the TRR130-P16 and TRR241-B03 (AR)

The Leibniz Association (Leibniz Collaborative Excellence, TargArt) (MFM)

COLCIENCIAS scholarship No. 727, 2015 (HRA)

## Author contributions

The concept of the study was developed by HRA, ES, ACL, ALS and TD.

Patient’s samples were collected by KB, FH, MC, UW, ES.

Data were obtained by FSZ, ALS, HRA, HS, BJ, KL, GMG, MFG, MFM, AP.

Data were analyzed by HR, ES, ALS, MFG, MFM and ACL, PD, FH.

The theoretical framework was developed by TD, ES, ALS and HR.

The work was supervised by ACL, KK, KUE, GB, KB and TD.

All authors developed, read, and approved the current manuscript.

## Data and materials availability

All data are available in the main text or the supplementary materials.

## Figures

**Figure S1: Nucleocapside ELISA among all donors.**Humoral immune response against SARS-CoV-2 was assessed by Euroimmune ELISA for nucleocapside in HC (n=25), DP (n=44) and KTR (n=40) 7±2 days after 2nd vaccination with BNT162b2. Kruskal-Wallis with Dunn’s post-test. *p<0.05, **p<0.01, ***p<0.001, ****p<0.0001.

**Figure S2: Subset distribution of RBD+ B cells of HC, KTR and DP.**Frequencies **(A)** and absolute numbers **(B)** of RBD+ plasmablasts (gating Figure 1A) naïve, pre-switch memory, post-switch and double negative B cells according to CD27/IgD in mature B cells (gating Figure 1A) in 7±2 days after 2nd vaccination in HC, DP and KTR.

**Figure S3: Expression of surface markers in 4 KTR by Cellular Indexing of Transcriptomes and Epitopes by Sequencing (CITE-seq).**UMAP representation of the expression levels of CD4, CD8, CD20, CD25, CD45RO, CD45RA and CD127 surface markers from 3 non-responders and 1 responder KTR 7±2 days after boost vaccination.

