## Supplementary figures and images for "Impaired antigen-specific memory B cell and plasma cell responses including lack of specific IgG upon SARS-CoV-2 BNT162b2 vaccination among Kidney Transplant and Dialysis patients"

### Supplementary Figure1

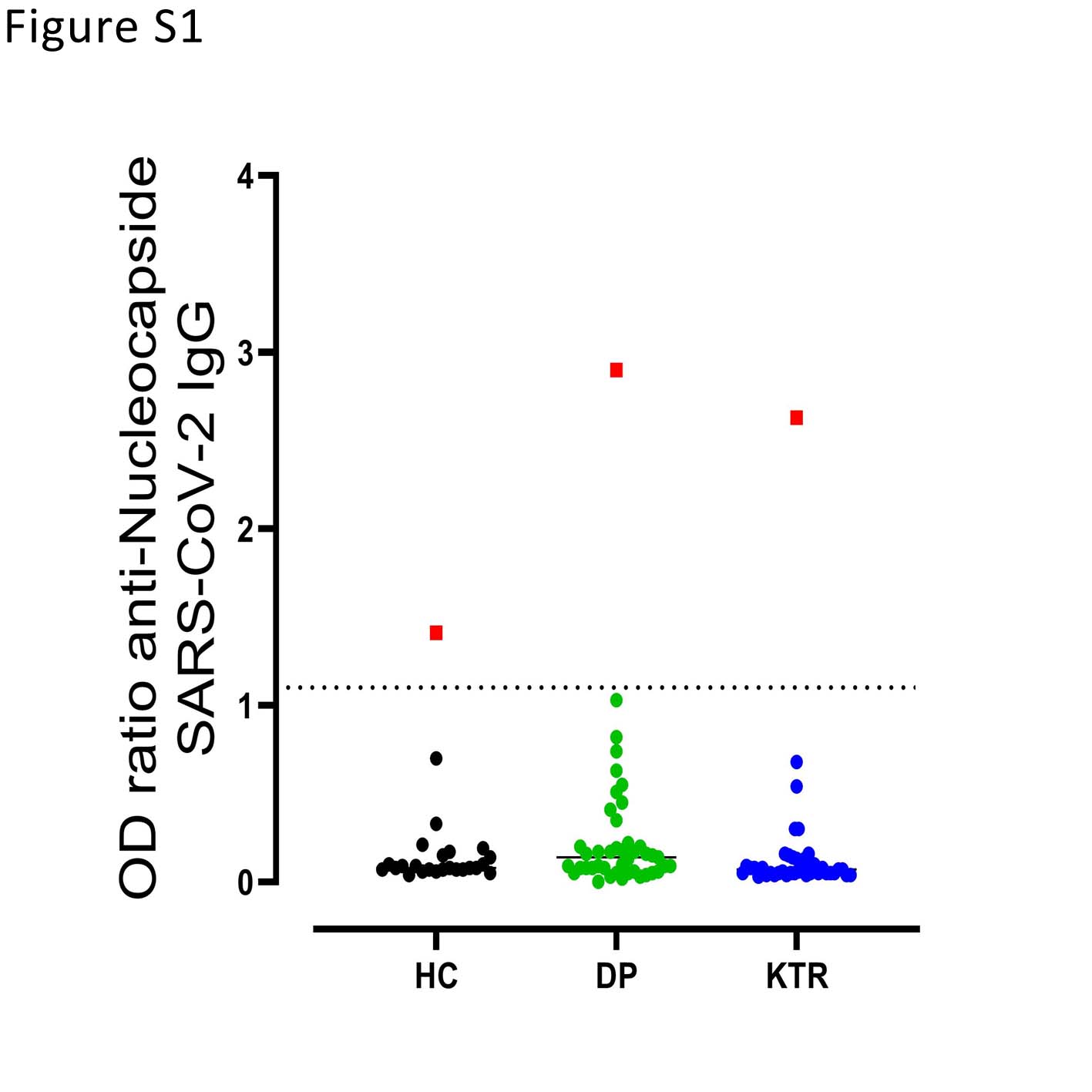

### Supplementary Figure2

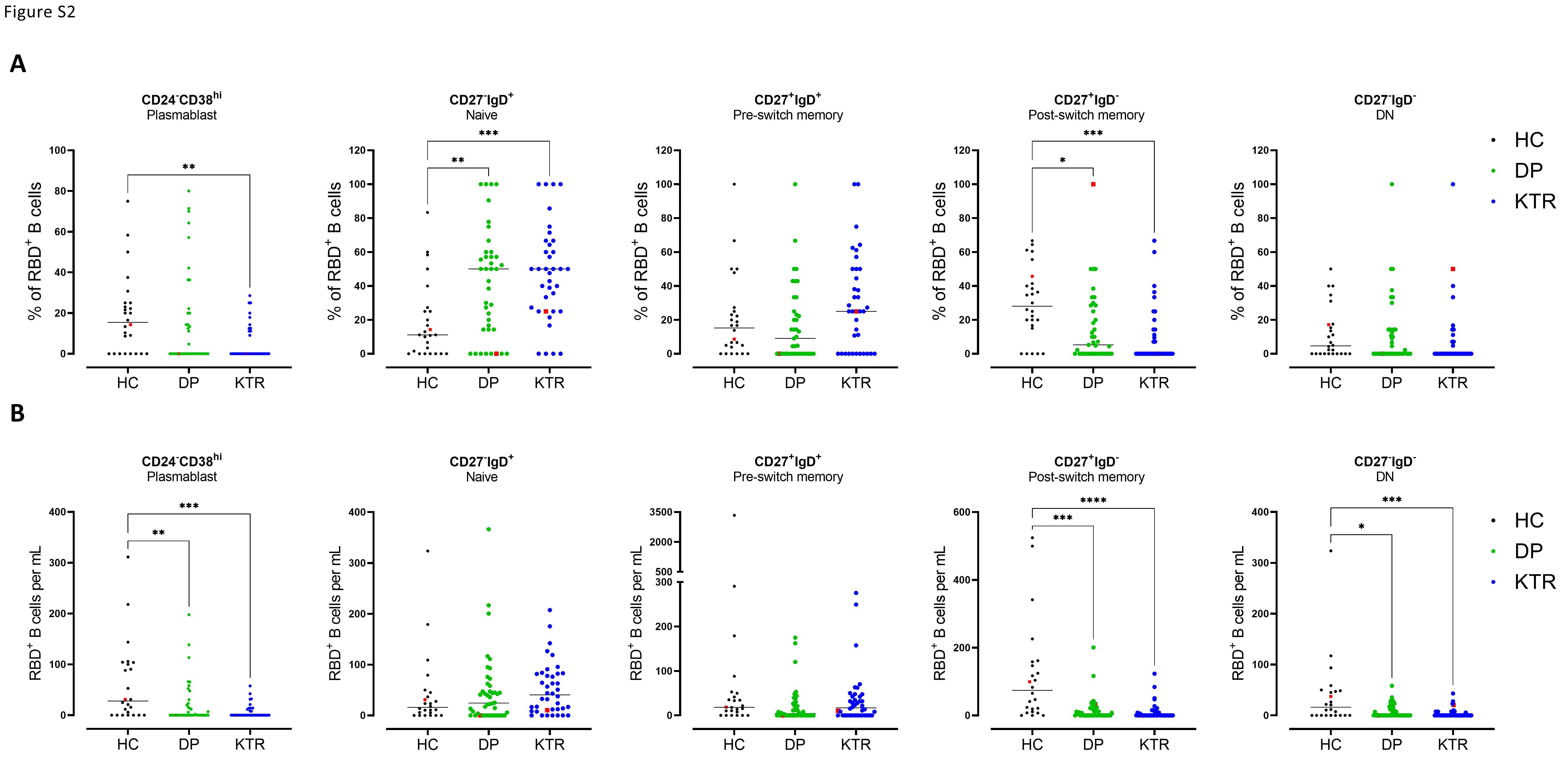

### Supplementary Figure3

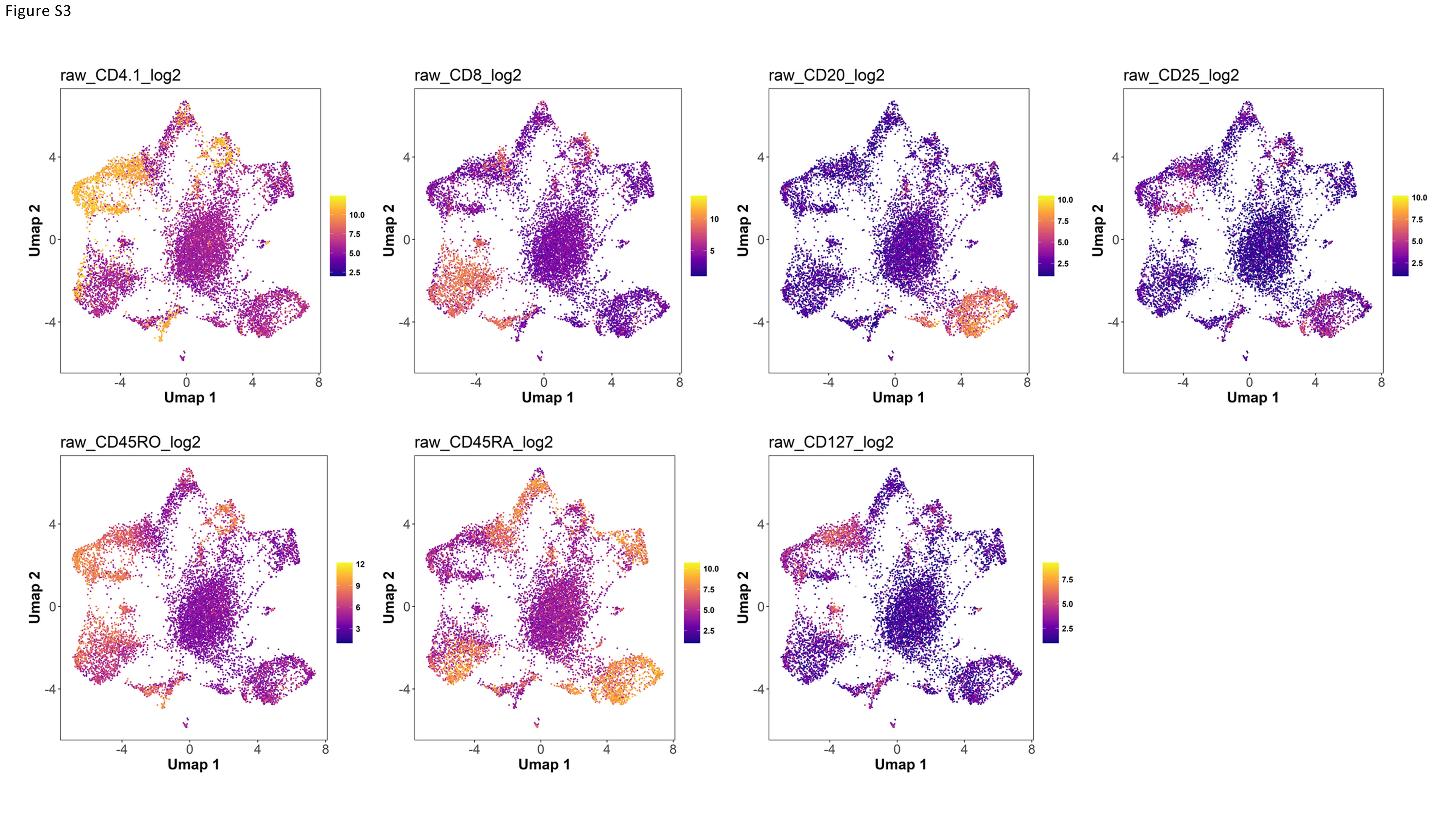
